# Rats and the COVID-19 pandemic: Early data on the global emergence of rats in response to social distancing

**DOI:** 10.1101/2020.07.05.20146779

**Authors:** Michael H. Parsons, Yasushi Kiyokawa, Jonathan L. Richardson, Rafal Stryjek, Kaylee A. Byers, Chelsea G. Himsworth, Robert M. Corrigan, Michael A Deutsch, Masato Ootaki, Tsutomu Tanikawa, Faith E. Parsons, Jason Munshi-South

## Abstract

Following widespread closures of food-related businesses due to efforts to curtail the spread of SARS-CoV-2, public health authorities reported increased sightings of rats in close vicinity of people. Because rats vector a number of pathogens transmissible to people, changes in their behavior has consequences for human health risks. To determine the extent of how stay-at-home measures influenced patterns of rat sightings we: 1) examined the number of rat-related public service requests before and during the period of lockdown in New York City (NYC) and Tokyo, Japan; 2) examined reports made in proximity to closed food service establishments in NYC; and 3) surveyed pest control companies in the United States, Canada, Japan, and Poland. During the month following lockdown, the overall number of reports decreased by 30% in NYC, while increasing 24% in Tokyo. However, new hotspots of 311 calls were observed in proximity of closed food service establishments in NYC; and there was a consistent positive association between kernel density estimates of food service establishments and location of 311 calls (*r* = 0.33 to 0.45). Similarly, more reports were observed in the restaurant-dense eastern side of Tokyo. Changes in clientele for pest control companies varied geographically, with 37% of pest-management companies surveyed in North America reporting 50-100% of their post-lockdown rat-related requests coming from new clients. In Warsaw, where there are no clusters of restaurants in densely-populated areas, there were no changes. In Tokyo, there were no changes in clients. We conclude that changes in public service calls are region-specific and localized, with increases in rat sightings more likely near restaurant-dense regions. Pest control companies surveyed in North America either lost much of their business or shifted clientele from old to new locations. We discuss possible mitigation measures including ramping up pest control during re-opening of food-related establishments and the need for citywide rodent surveillance and disease monitoring.

## Introduction

Urban rats (*Rattus spp*.) are global commensal organisms that depend on humans for food and harborage. Thus, shifts in human behavior, such as occurs following natural catastrophe or pandemics, will have pronounced effects on nearby rat populations. This effect has been hypothesized after social distancing and business closures were broadly enacted in Spring 2020 to limit the spread of SARS-CoV-2. Shortly thereafter, local governments and public health authorities around the world began reporting that closures of restaurants and food-related venues have coincided with reports of mass sightings of rats^1-3^. These sightings include reports of aggressive behaviors during daylight hours and in close proximity to people^4,5^ with some rats consuming conspecifics (e.g. muricide or cannibalism;^6^. Given the heightened state of anxiety among people affected by the social distancing mandates^7,8^, these sightings could also be a reflection of human sensitivity toward rats as indicators of poor sanitation and disease^9,10^. Further, due to the increased use of social media during the pandemic^11,12^ and the rate at which ‘panic’ spreads as a social contagion globally^13^, predisposition or cognitive bias could lead to increased reports. These reports, while broadly circulated in the popular media, have yet to be reviewed or confirmed through research or surveillance.

The potential mass movements of rats into new areas may negatively impact human society^14,15^. Rats are known to transmit many types of disease^14,16,17^ and are associated with billions of dollars in losses of food annually^18,19^.They are commonly thought to have killed more humans than all wars combined^20,21^, and these fearful perceptions have caused an intense fear of rats in many cities^22^. The mere presence of rats is enough to cause harm to mental well-being, particularly in low socio-economic status areas^10,23^. But despite these harmful consequences, there are no validated methods to quantify rat movements irrespective of human reports^24-26^. Further, there are no routine disease surveillance programs to detect changes in pathogens that rats may be transmitting^14^. Thus, cities are limited to the reporting of rat sightings to determine spatial risks of rodent-borne disease and where abatement programs should be concentrated^27,28^.

While cities need new mechanisms to monitor rat populations, it is important to understand the relationship between human behavior and rat activity. Changes in the activity of either species may have profound implications for the other^22,23,27^. In temperate regions where seasonal changes are prominent, rats and humans concurrently increase activity during Spring and Summer, which results in predictable increases in rat sightings. For instance, reports of rat complaints in Chicago over a ten-year period, 2008-2018, demonstrated a particularly strong rise from winter to spring, then peaks each summer and decreases again from fall to winter^29^. These seasonal fluctuations in complaints can be explained by both decreased activity of rats as they retreat deep within their burrows in colder months^30,31^ and the reduction in human outdoor activity in winter^32,33^. Human reporting of rat sightings can also be impacted by contextual bias and cognitive bias^34^. For instance, rats observed in new environments are more likely to be noticed and reported than rats in areas where they have previously been established. Further, a single rat sighting in a public area might generate more complaints compared to those in less visible, or private, areas^15^. Lastly, the bandwagon effect^35^ is a type of predisposition that causes people who have knowledge of a widely-communicated event such as the global media reports on rats, to have an increased awareness of rats, and thus, be more likely to report them.

Because humans have undergone profound changes resulting from social distancing and isolation and because commensal rat populations are commensal with humans, rat populations are almost certainly affected, however the nuances of these impacts remain unknown. Our intentions for this paper are to provide data to help us understand to what extent changes in human reporting behavior and/or wild rat ecology were responsible for the widely-reported phenomenon. In colder climates, such as found in New York City, we sought to identify observable shifts in public service reports (e.g., 311 calls) independent of seasonal effects^29^. Calls to 311 are free to report. Calls reported to pest management companies^36^ on the other hand, convey an added sense of urgency as they carry a monetary expense. Due to detailed, publicly available data for the city, we were also able to examine food availability for rats in the form of restaurants and food carts. In Tokyo we utilized public calls to the Tokyo Pest Control Association (TPCA) which are used as a first step prior to hiring an exterminator, along with cost-carrying reports made directly to private pest control companies. Neither Poland nor Canada have immediately accessible public service reports, but national pest management associations exist in each country, which we used to distribute surveys.

## Methods

We analyzed public service requests (311 calls) from NYC Open Data (https://opendata.cityofnewyork.us/) on May 3, 2020 for the observation period beginning January 1, 2014 and ending April 30, 2020, inclusive of the lockdown phase which began on March 23, 2020. New York City was considered an appropriate region for analysis due to its high human population density, and robust, regularly-updated and easily accessible reporting data. Reports are made when people file a complaint through the city’s 311 via phone, website or smartphone app. The selected interval was chosen because it allows a before-and-after treatment analysis to be considered alongside annual seasonal changes. Rodent-related 311 calls are classified into 5 categories: signs of rodents, conditions attracting rodents, rat sighting, mouse sighting, and rodent bites. For this paper, we limited our analysis to the “rat sighting” category.

In Tokyo, the Japanese Government requested self-isolation on Feb. 16 and then declared a state of emergency on Apr. 7. Therefore, we analyzed the number of phone calls to the TPCA from January through April from 2015 through 2020, which was provided by the TPCA. The TPCA is a non-profit public corporation established by the Ministry of Health & Welfare of the Japanese Government. As a public interest incorporated association, they provide services and referrals to the general public and also have access to 105 pest-management companies. Complaints to the TPCA and subsequent consultation are free of charge, other than cost of a local call. The TPCA introduces its members to pest control operations when necessary. However, people are still responsible to decide whether or not they will contract with a pest-management companies.

Pest control interventions offer a means to determine prevalence of rats^36^. Thus, we also created an industry survey with the help of industry professionals (Appendix A). The survey covered at least a 30-day period from lockdown to post-lockdown for each nation. Three primary questions were posed: 1) whether there had been changes in overall rat-related calls from customers; 2) what approximate proportion of post-lockdown customers were new customers (which may account for customers in new residences not previously infested); and 3) whether the number of new customers for rat jobs was different from the number of new customers during the same period the previous year. Respondents were not required to disclose personally identifiable information. The survey was distributed through pest control channels (CPCA) throughout Canada on May 5, 2020 and throughout the United States via Pest Control Magazine ^4^ as well as via social media through Twitter and LinkedIn. The survey was translated in Japanese and distributed to the 105 TPCA member pest-management companies on May 11, 2020. Because Japan has a large number of roof rats, the survey additionally asked about the proportion of roof rats, brown rats, and house mice being reported. The survey was also translated in Polish for dissemination via pest management channels, but due to the low numbers of roof rats in Poland, there was no request for the species of rat being reported.

### Statistics

#### Phone calls

We used one-way ANOVA to identify overall differences in average daily rat sightings per month across all years in the observation period, and Tukey’s HSD test for pairwise comparison between months. We then used two-way ANOVA to test the association between the daily number of rat sightings in the months of March and April during the previous 6 years without social distancing, 2014 to 2019, and during the social distancing phase in year 2020. We used Tukey’s HSD test to conduct pairwise comparisons between the periods without social distancing and the period with social distancing for the month of March, and similarly for the month of April.

The Tokyo metropolitan area includes 23 wards, 26 cities, 3 towns, and 1 village. Among them, the “accommodation, eating, and drinking services” (79.9% in 2016, based on Tokyo Metropolitan Government website) and population (69.2% in 2020, based on Tokyo Metropolitan Government website) were heavily biased toward the eastern 23 wards area (**Fig.2**). Therefore, we focused on the data in 23 wards. From 2018 to 2020, the number of calls in the target month was compared with the average of previous 3 years in the same month. When the number of calls in 2020 was increased or decreased by more than 2 standard deviation as compared to the previous 3 years, the number was classified as “increased” or “decreased”, respectively. The differences in the number of wards showing “increased”, “decreased,” and “no change” among 2018, 2019, and 2020 were analyzed by Fisher’s exact test.

#### Spatial analysis

To investigate changes in the spatial distribution of rats in NYC over the study period, we mapped 311 rat calls in GIS and applied hot spot analysis. Both ArcMap 10.6 and QGIS 3.10 were used for these analyses. We first divided 311 calls for NYC by month and year and imported these as separate data layers in GIS. We generated a time series of 311 rat calls in QGIS by creating a heatmap and working within the Time Manager plugin. We then conducted an optimized hot spot analysis in ArcMap using the Mapping Clusters toolbox. This analysis evaluates areas with more or less point occurrences than expected at random, and applies a Getis-Ord G*i* statistic to assess significant deviations from a random distribution. We used the NYC borough outline as the bounding polygon for this analysis, and a fishnet pattern was used to generate sampling polygons across the study area, as recommended for this tool. Lastly, we created kernel density raster layers of both the food service establishments (using NYC Open Data on inspections) and rat 311 calls by month, and used the Band Collection Statistics toolbox to calculate correlation coefficients between each raster dataset.

#### Survey

Descriptive statistics were used to report survey findings. In Tokyo, a two-way repeated ANOVA was used to analyze the proportion of rodents. Planned comparisons were conducted using paired t test to clarify the changes caused by a state of emergency and Tukey HSD test to clarify the predominant species before and after a state of emergency.

## Results

### 311 calls

In New York City, one-way ANOVA showed a significant difference in daily rat sightings among the 12 months throughout the observation period (F=106.5; *P*<0.001). Tukey’s HSD tests indicated that, from 2014 to 2018, there are five groups of months with similar number of rat sightings that appear to follow a seasonal trend. Specifically, the lowest number of daily rat sightings were lowest in the winter burrowing periods (December and January) while the highest were in summer (July through August). April sightings were significantly different from all the other months, while March and November were similar (**Fig 1A**).

**Figure 1.**
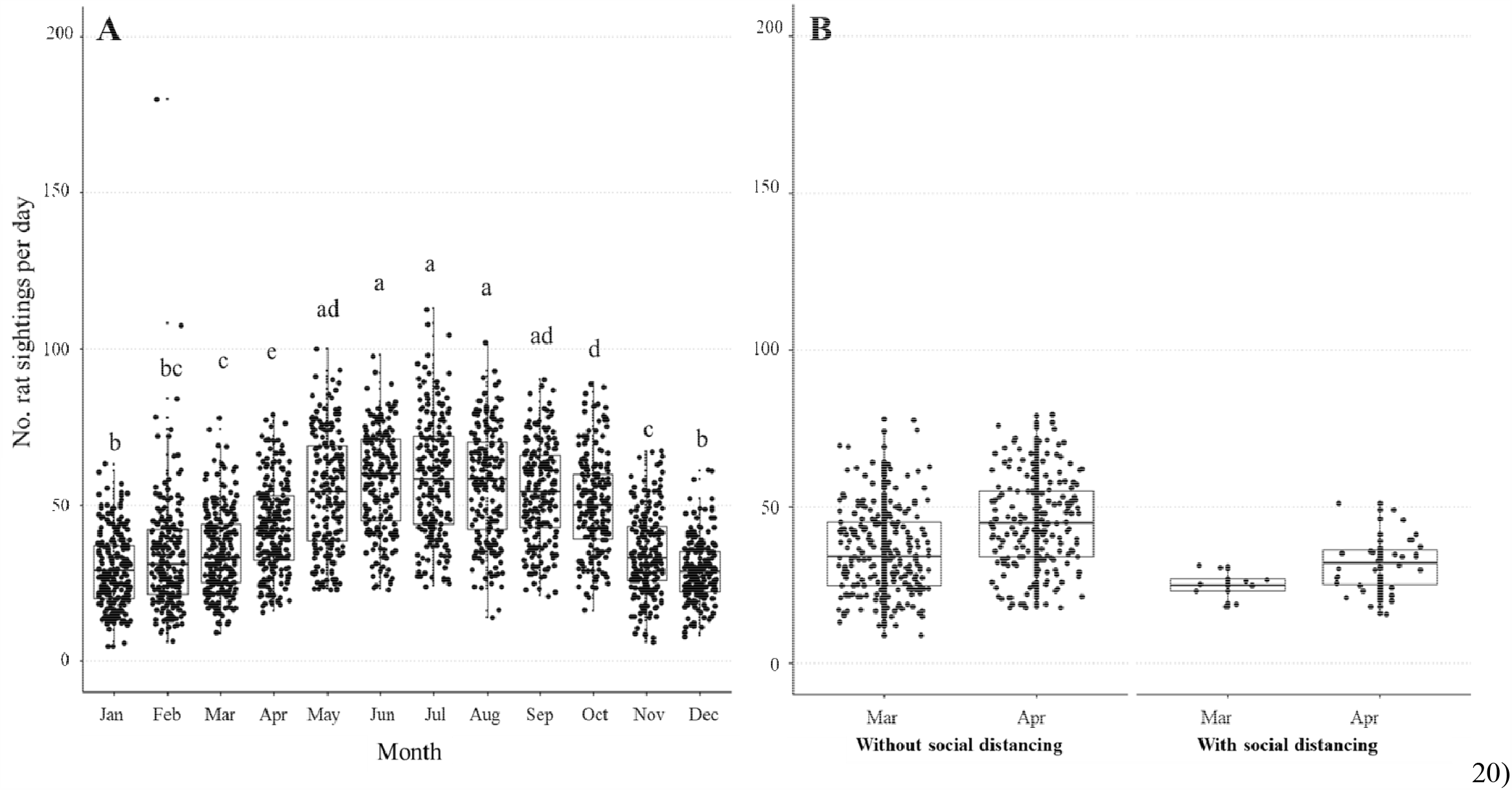
Right, the association between number of reported rat sightings per day by month with reference to the social distancing period of March and April 2014 to 2019 compared with the same two months during the social distancing period, year 2020. Left, distribution of daily rat sightings per month during the observation period (January 2014 to April 20) using data from NYC Open Data.

Two-way ANOVA showed a significant association between the average number of rat sightings for the months of March and April during the years without social distancing (2014 to 2019) compared to the current year (2020) where social distancing was enforced (**Fig 1B**, F_1_=35.4; *P*<0.001). For the month of March, the early part of the COVID-19 lockdown period where social distancing was enforced, there was no significant difference in the average number of sightings during the social distancing phase 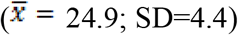 as compared to March in the prior 6 years when there was no social distancing 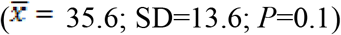. However, for the month of April, the first full month of isolation orders in NYC, the number of rat sightings during the social distancing period 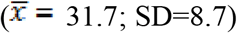 was significantly lower compared to April in the prior 6 years without social distancing 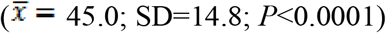. Although the number of rat sightings in April 2020 was significantly lower compared to the same month in prior years, the figure shows an increasing trend in the reported rat sightings from March to April 2020 (consistent with the trend shown in **Fig. 1A)** however, the difference is not statistically significant (*P*=0.560).

### Spatial relationship with food service establishments

There was a positive correlation between the kernel density estimates of food service establishments and location of 311 calls in all of the months assessed (Fig. 4; **Supplemental Table 1**). However, the spatial association between food service establishments and rat 311 locations did increase between the pre- and post-lockdown periods (*r* of 0.35–0.36 in January/February 2020 compared to 0.44–0.45 in March/April 2020). Additionally, the Getis-Ord analysis identified hotspots of 311 complaints within NYC, which changed only modestly before and after the COVID lockdown period (**Fig. 4**). The correlation analysis also showed a high correlation among 311 calls throughout the 3 years (*r* = 0.83 to 0.95), which indicates that the locations of the rodent complaints are generally consistent across the years and months analyzed. There were no detectable hotspots in areas away from clusters of food service establishments (**Supplemental Fig. 1**).

#### Calls to TPCA (Tokyo)

Within the 23 wards of interest in Tokyo, the total number of calls from January through April. were 224 (2015), 296 (2016), 293 (2017), 299 (2018), 314 (2019), and 365 (2020) which ranged from 47.3% to 54.2% of calls in the Tokyo Metropolis. In the 23 wards, the total number of phone calls was increased in Jan. and Apr. 2020, but not changed in February and March. The patterns of changes in Tokyo were shown in **Fig 2**. In January, 5 of 6 changes (4 increased and 1 decreased), 2 of 4 changes (2 increased), and 1 of 4 changes (1 decreased) occurred in the 23 wards in 2018, 2019, and 2020, respectively. Fisher’s exact test revealed that the changes in Tokyo (*P* = 0.55) and in 23 wards (*P* = 0.16) were not different among three years. In February, 5 of 5 changes (3 increased and 2 decreased), 5 of 7 changes (5 increased), and 6 of 10 changes (5 increased and 1 decreased) occurred in the 23 wards in 2018, 2019, and 2020, respectively. Fisher’s exact test revealed that the changes in Tokyo (*P* = 0.23) and in 23 wards (*P* = 0.72) were not different among three years. In March, 3 of 5 changes (2 increased and 1 decreased), 2 of 4 changes (2 decreased), and 4 of 7 changes (4 increased) occurred in the 23 wards in 2018, 2019, and 2020, respectively. Fisher’s exact test revealed that the changes in Tokyo (*P* = 0.08), but not in 23 wards (*P* = 0.16), were significantly different among three years. In April, no change occurred in 2018. While 1 of 2 changes occurred in the 23 wards in 2019, 5 of 7 changes occurred in 2020 (5 increased). Fisher’s exact test revealed that the changes in Tokyo (*P* < 0.05) and in 23 wards (*P* < 0.05) were different among three years.

**Figure 2.**
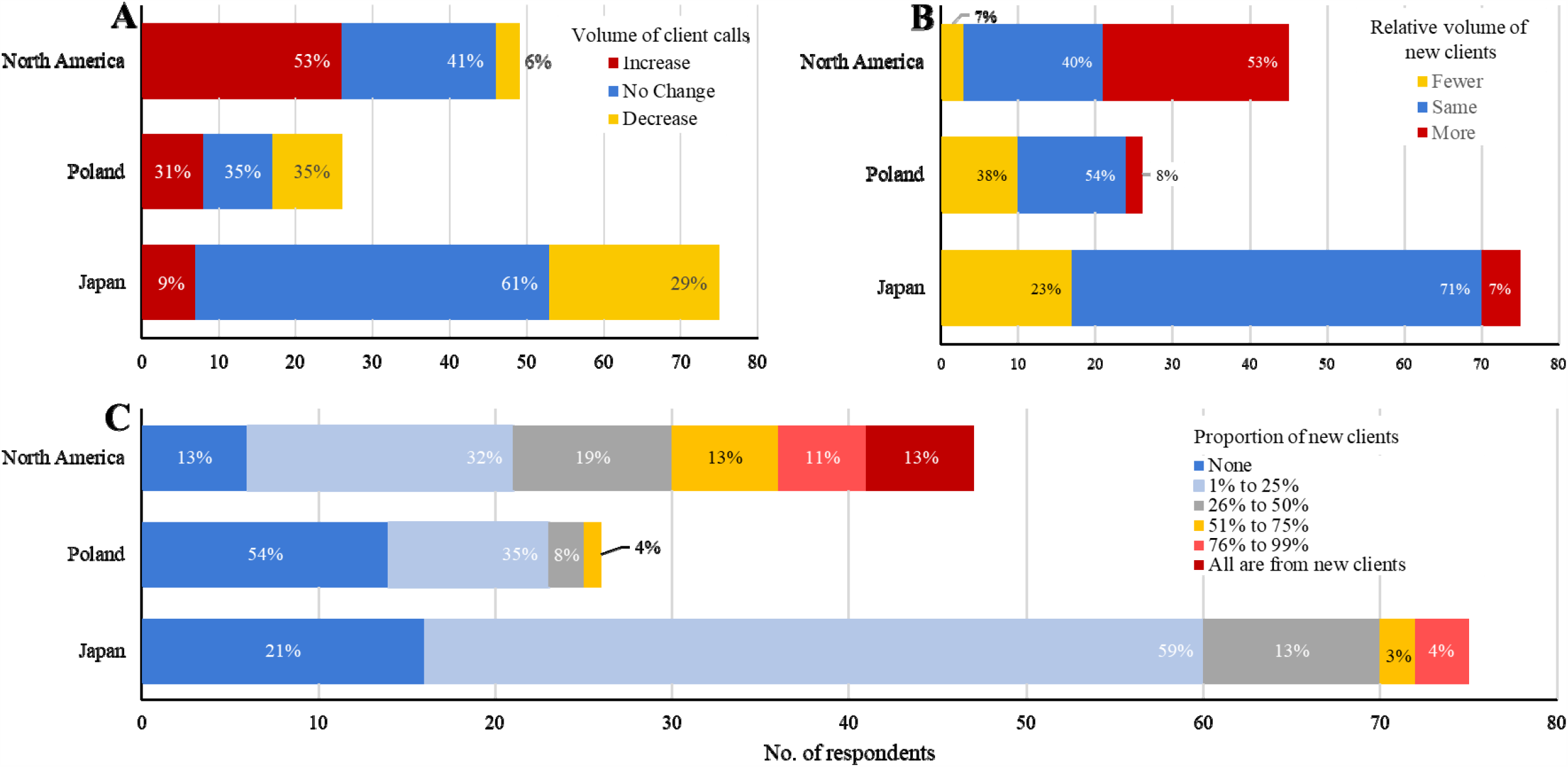
New and old clients. Survey responses from 53 pest control companies in North America, 26 in Poland, and 75 in 23 wards of Tokyo, Japan. Top right, number of all rat-related calls/jobs from new clients. Top left, relative volume of overall rat-related calls/jobs before and after the lockdown period. Bottom, proportion of new client calls due to rat-related jobs.

### Survey Data

There were 50 respondents from North America, 26 from Poland, 85 from Tokyo, and one each from Argentina, India, and Malaysia. Of the 50 respondents in North America, one responded “*prefer not to answer*” to the first question on whether there has been changes in number of rat-related calls from customers, 3 to the second question on whether the majority of post-lockdown customers were new customers, and 5 to the third question on whether the number of new customers for rat jobs was different to the number of new customers during the same period the previous year. The responses from Argentina, India and Malaysia were excluded from the analysis, and responses of “*prefer not to answer*” were excluded on a per question basis. Data from pest management companies from the US and Canada were combined as there was a similar pattern in responses. The 50 respondents were from 47 cities across North America. Given the number of cities represented, compared to the total number of respondents, we assume that we received one response per pest management company.

Of 85 respondents in Tokyo, 82 responses were included as all questions were answered clearly. The 82 respondents covered of 29 of 53 areas of Tokyo and all 23 wards. We focused on the 75 respondents from the 23 wards, because these wards, along with Tokyo, function as a single urban entity with respect to public services such as rat complaints. The number of questions answered ranged from 1 to 9 in each area. All respondents from Poland answered all three questions and were all included in the analysis.

### Volume of rat-related calls or jobs

More than half of the respondents in North America (55%, **Fig. 3A**) indicated that they received an increase in the volume of rat-related calls or jobs, while only 9% of responses from the 23 wards reported an increase. In Tokyo, the majority of the responses from the 23 wards (61%) indicated that there was no change in the volume of calls. And in Poland, the distribution was very similar between those who experienced an increase, a decrease and no change in the overall volume of rat-related calls or jobs.

**Figure 3.**
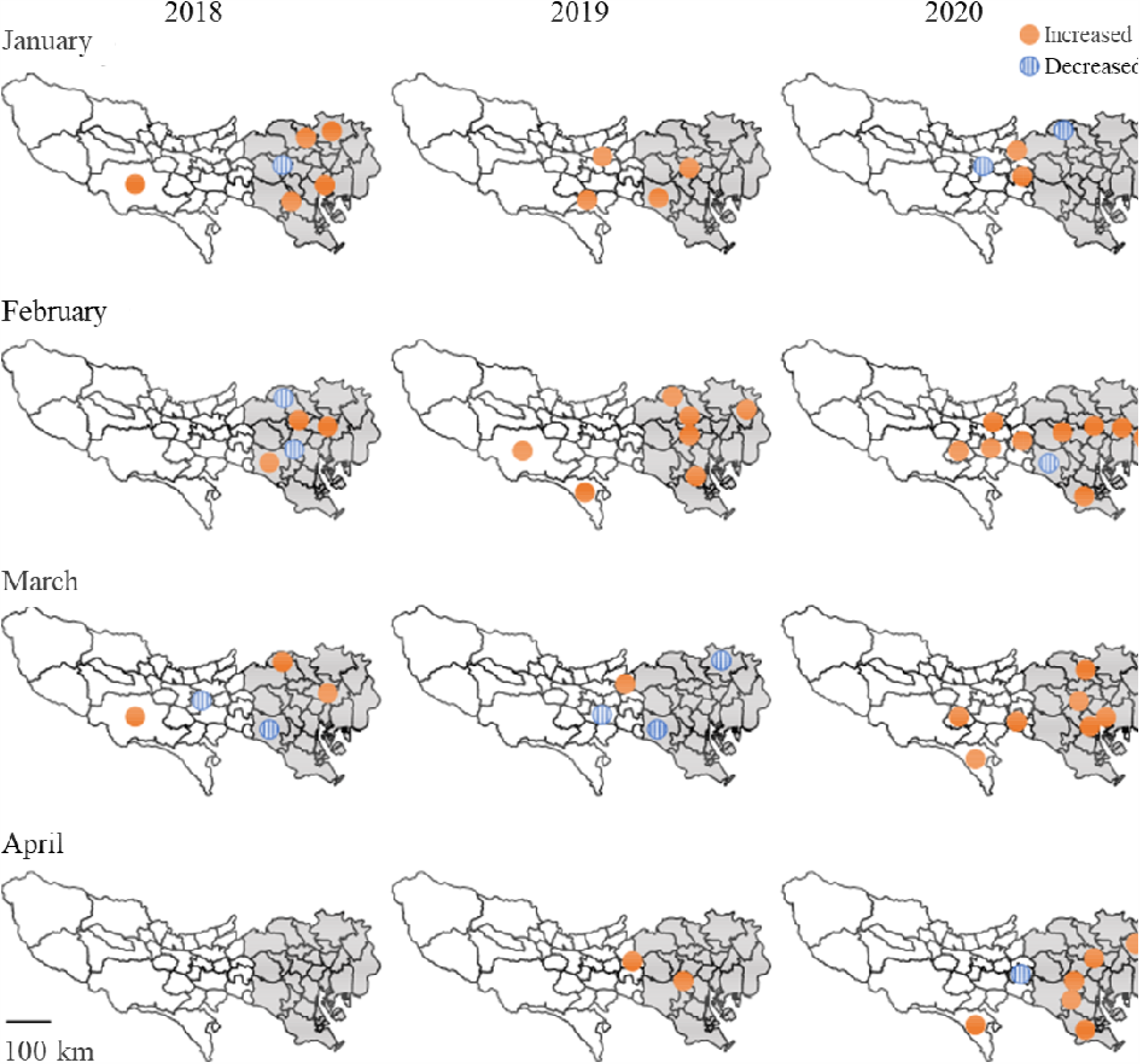
The changes in service calls to Tokyo Pest Control Association over period from January through April from 2018 to 2020. The grey areas indicate the 23 wards.

**Figure 4.**
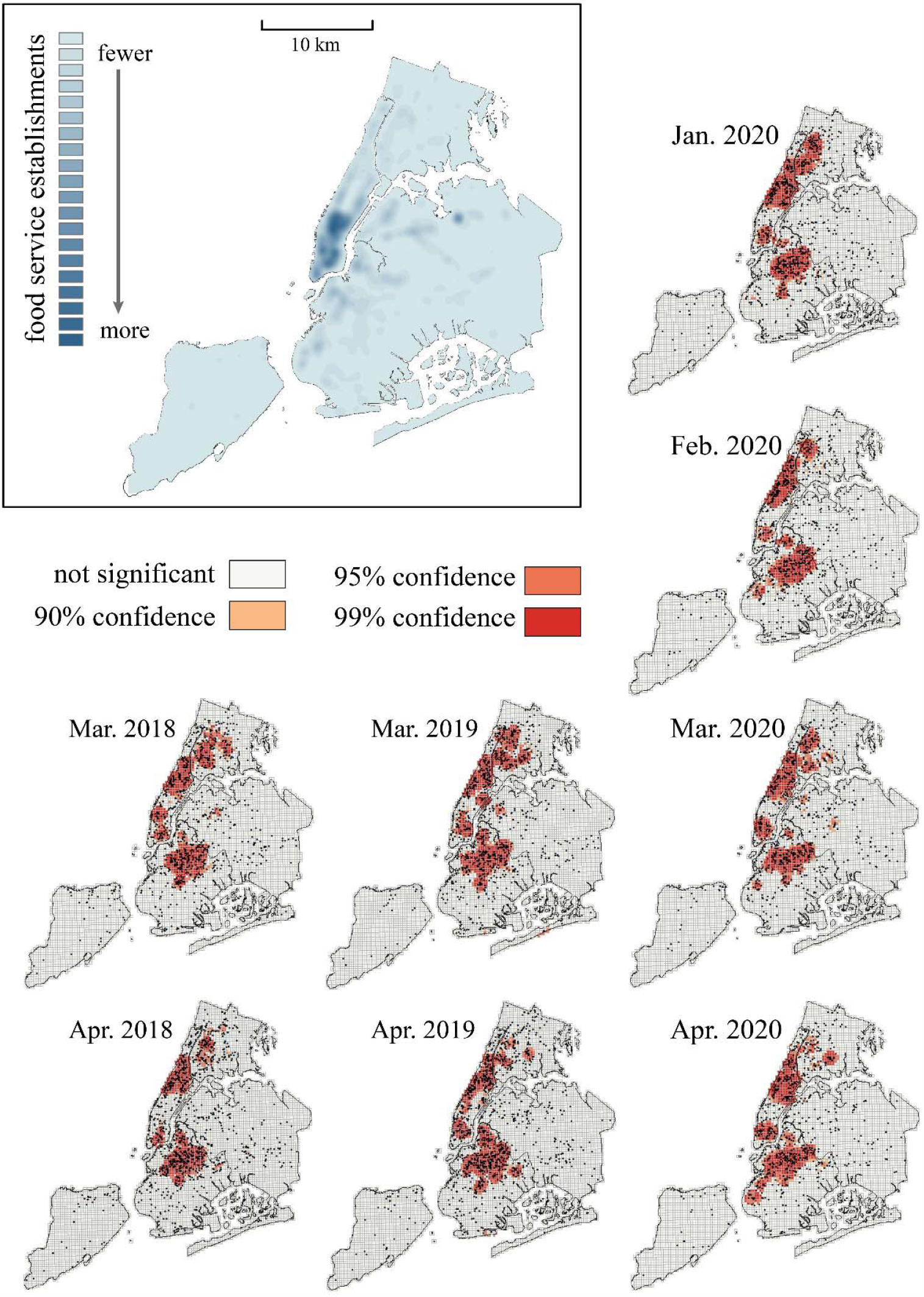
Kernel density of food service establishments (top left) and results of optimized hotspot analysis (Getis-Ord G*i*\*) of 311 calls in New York City, USA (right and bottom).

### Relative volume of rat-related calls or jobs from new clients

More than half of the respondents from North America (**Fig. 3B**, 53%) reported that they have more calls from new clients for rat-related jobs as compared to the previous year. This is in contrast with the responses from Poland (8%) and Tokyo (7%) where the majority of the responses indicated that the relative volume was the same as compared to the same time in the previous year (54% and 71%, respectively).

### Proportion of rat-related jobs from new clients

When asked how many of the rat-related calls or jobs were from new clients, 13% of respondents in North America indicated that all of their rat-related jobs were for new clients, while none of the respondents from Tokyo or Poland selected this response (**Fig. 3C**). More than half of the respondents in Poland (54%) answered that none of their rat-related jobs were from new clients, while only 21% in Tokyo and 13% in North America selected the same answer. Most of the respondents from Tokyo (60%) indicated that the proportion of rat-related jobs from new clients were very low, 1% to 25%, while the same response was obtained from one-third of the respondents from both Poland (35%) and North America (32%).

Among the 24 respondents in North America who experienced an increase in rat-related calls or jobs, 21 (88%) indicated that they have acquired more new clients for rat-related jobs compared to the same time last year, while the remaining 3 (12%) responded that the relative volume is about the same.

### Proportion of rodents in Tokyo

Before declaration of a state of emergency, the proportion of brown rats, roof rats, and house mice in Tokyo 23 wards was 20.5 ± 1.7 %, 78.9 ± 1.8 %, and 0.7 ± 0.3 %, respectively. The proportion changed to 21.6 ± 2.0 %, 77.8 ± 2.1 %, 0.7 ± 0.3 %, respectively, after the declaration. The proportion was significantly affected by the type of rodents (*F*(2,225) = 748, *P* < 0.01), but not by the declaration of a state of emergency (*F*(1,225) = 0, *P* = 1). Interaction of these two factors was not significant (*F*(2,225)=1.32, *P* = 0.27).

A planned comparison by paired t test did not find significant changes by the declaration of a state of emergency (brown rats: *t*(75) = 0.94, *P* = 0.35; roof rats: *t*(75) = -0.94, *P* = 0.35; house mice: impossible to assess because the majority was 0). A planned comparison by Tukey’s HSD test revealed that the proportion of roof rats was significantly higher than those of brown rats (*P* < 0.01) and house mouse (*P* < 0.01) both before and after a declaration of a state of emergency. The proportion of brown rats was higher than those of house mouse both before and after a declaration of a state of emergency (*P* < 0.01).

## Discussion

There are currently no validated measures to identify rat behaviors or movements in the city despite their importance to human health risks. However, two key pieces of evidence suggest that changes in human reporting behavior have to some extent been precipitated by changes in rat movements. First, we have found a surprisingly high association between the location of rodent complaints and food service establishments and; secondly, we provide evidence that these ‘hot spots’ of rat sightings have shifted during the pandemic. Had these changes in rat reports been a result of human cognitive bias^34^ and/or hypervigilance due to ‘the bandwagon effect’^35^, then changes would have likely been unrelated to food businesses and more geographically uniform. Instead, changes in public service calls and requests for pest control were region-specific and localized, with increases in rat sightings in New York City (NYC) and Tokyo more likely to be reported within, or near, restaurant-dense regions. In Warsaw, where there are no clusters of restaurants within densely-populated areas, there were no changes.

The fact that calls decreased in NYC was unanticipated because institutions that reported rat sightings^1-3^ implied there were many more sightings than usual. The decrease in sightings was further surprising because the period of lockdown coincided with the spring season when rat sightings typically increase as rats emerge from underground burrows^29^. Yet, the post-pandemic reports moved in the opposite direction from the previous 6 years, with a 30% decrease in volume of 311 calls in NYC. Conversely, in Tokyo, we were surprised that while calls actually did increase by 23% as compared to 5 previous years, there were no differences between surveys administered before and after the lockdown period. However, within North America, the shift in clientele was striking. More than one-third of pest management companies surveyed reported 50-100% of their post-lockdown jobs were from new clients, while six companies reported that 100% of their client-base had changed from old to new clients. Within the pest management industry, such pronounced changes in rat populations following catastrophes are expected^37,38^.

Some possible explanations for the geographic variability in outcomes relate to differences in rat control, human social behavior and the ecological differences between the primary species of rats being reported. In NYC, the lockdown was accompanied by the widespread adoption of social distancing practice. The highest concentrations of sightings typically occur near subway lines and recreational public spaces^39^. However, in April 2020, the majority of residents were at home social distancing and many had left the city (e.g., counter-urbanization^37^). Additionally, due to the abrupt reduction in available food, many rat populations had temporarily decreased due to stress and competition. Another possible explanation is that people who found an infestation near their residences may have contacted pest control directly rather than reporting to 311. This finding makes sense because prior to restaurant closures a single rat might be seen by hundreds of patrons and reported. During business closures the opposite might be expected to occur, where many rats in a single residence might elicit only a single call—likely to pest control for immediate response and not 311.

In Tokyo, it was surprising that the increased phone calls to TPCA were not reflected in increased business for pest-management companies. This might be because the two species of rats present in Tokyo were affected differently by the lockdown as roof rats were responsible for 78% of complaints registered in the surveys, while Norway rats made up only 20%. In addition to being adapted for warmer climates, roof rats are more difficult to spot; they are smaller, arboreal, make nests inside of ceilings or roofs, and migrate shorter distances. Pest control in Tokyo is also mandatory in large buildings and thus roof rats are most likely found inside smaller buildings such as restaurants and bars. Because restaurants and bars remained open for ‘take-away’ meals, roof rats could continue to feed on food stocks, grease vats, and oil stains. Calls regarding this species would have not been impacted greatly by the lockdown.

In contrast, the eastern side of Tokyo, where calls increased, is composed of the downtown area where restaurants are more concentrated. In this area, large amounts of garbage are placed street-side at midnight where Norway rats can consume garbage *ad libitum* until collected the following morning. Thus, it is highly probable that roaming by Norway rats increased the overall number of phone calls to TCPA, but was not perceived as important enough to hire a private pest control specialist. We should also note that in Tokyo most pest-management companies contract with restaurants and bars in one-year installments. The businesses that utilize these contracts for the majority of their clientele would not be expected to be affected much by the declaration. Specifically, if the number of one-year contracts had been much larger than new contracts, then surveys of pest control companies would have failed to reveal the changes caused by the declaration. In any case, we found that pest-management companies were not impacted as prominently as NYC. Poland does not have a public service request system; however, they do enforce mandatory pest control with fines for offenders with rats on their properties. Given that they do not have clusters of restaurant-dense areas in heavily-populated areas, it is reasonable that there were no changes in pest control needs before or after lockdown.

If large numbers of rats are migrating in some areas or regions—as appears to be supported by our findings—then certain characteristics of immigrant rat populations may change^40^. For instance, if formerly insular rat populations^41^ come into increased contact with one another due to migration, then mating opportunities could occur between colonies that do not typically interbreed. Additionally, if large numbers of rats concurrently migrate, then a greater variety of individual genotypes within populations would have moved significant distances while under intense selective pressure^40^. It is reasonable then to assume there is some influence on the genetic profile of future urban rat populations^42,43^. While stressed rat populations may initially decrease due to loss of food and increased competition and muricide, rats breed rapidly, and should rapidly re-establish population equilibrium^44^. Impacts on human populations are less clear.

Rodents are considered unlikely candidates to be infected with or transmit SARS-CoV-2^45^. Only two rodent species, the atypical 5 kg giant bamboo rat (*Rhizomys sinensis*)^46^, and the golden Syrian hamster (*Mesocricetus auratus*)^47^ have been associated with the COVID-19 virus.. However, stressed or wounded rodents are less likely to groom and keep their fur clean, and thus present a greater hazard to mechanically transmit contagious diseases without being infected themselves^48,49^. Additionally, in the case where rodents transmit pathogens that affect the immune system, such as Lyme disease, then those affected could be at increased risk of a weakened immune response to other pathogens^50,51^. These increased risks to human health support the urgent need for more robust data on rats in cities so that we can adequately track these changes and make informed decisions moving forward regarding management.

## Conclusions

Reports of increased sightings of urban rats near food service establishments suggest that lockdown procedures have resulted in increased rat movement, which could, in turn, alter the distribution of rats throughout the city. Because rats carry a number of pathogens that are transmissible to people, these changes may have potential negative impacts for humans. However, it is difficult to determine the extent to which social-distancing measures have impacted rat biology due to a general lack of systematic data collected on rat presence and abundance. To overcome these limitations, we combined 311 data, geographic data, and survey data to infer whether rat sightings have increased in relation to stay-at-home measures put in place during the COVID-19 pandemic. We demonstrate that following lock-down measures in NYC, that rat complaints decrease as compared to previous years. Further we show that changes in calls to pest control companies in North America, Japan, and Poland vary. Interestingly and importantly, we have demonstrated that that in North America there is a striking increase in the number of calls from new clientele, suggesting that stay-at-home measures have resulted in noticeable changes in rat presence.

Natural disasters typically result in temporary decreases in rat populations followed shortly by an uptick in populations^37,38^. Residents near food service establishments should therefore be prepared for local increases in rat populations during the late spring and early summer. Properties closest to restaurants should pay special attention to exclusion techniques and pest control to ward off the increased risks. General mitigation measures include public education regarding urban hygiene and widespread, systemic changes in how we manage our garbage^52^. We recommend that all commensal species^53^ be tested for their ability to harbor or transmit the virus, especially until a vaccine is developed. These assays could be particularly useful because rats frequent sewers where they come into contact with human wastes which were recently shown to contain heavy loads of the COVID-19^54,55^. Given the impact of rats on public health, and the challenges of exploiting human reports to learn about rats, there is a clear need for more consistent, rigorous, and standardized rat monitoring and disease-monitoring on a global scale.

## Data Availability

All raw data will be made freely available upon request. Additionally,

## Acknowledgements

We thank the Cornell University Extension Service (Matt Frye), Canadian Pest Management Association (Amy Cannon), Pest Control Technology (Brad Harbison), and National Pest Association Fairfax, Virginia (Jim Fredericks) for support and internal distribution of surveys. Prof. Stanislaw Ignatowicz helped distribute surveys in Poland.

---

**Supplemental Table 1.** Correlation matrix between the food service establishment (FSE) kernel density raster and monthly 311 calls kernel density raster in New York City, USA.

|  | FSE | Mar 2018 | Apr 2018 | Mar 2019 | Apr 2019 | Jan 2020 | Feb 2020 | Mar 2020 | Apr 2020 |
| --- | --- | --- | --- | --- | --- | --- | --- | --- | --- |
| **FSE** | 1 |  |  |  |  |  |  |  |  |
| **Mar 2018** | 0.40 | 1 |  |  |  |  |  |  |  |
| **Apr 2018** | 0.33 | 0.95 | 1 |  |  |  |  |  |  |
| **Mar 2019** | 0.42 | 0.88 | 0.89 | 1 |  |  |  |  |  |
| **Apr 2019** | 0.44 | 0.90 | 0.92 | 0.93 | 1 |  |  |  |  |
| **Jan 2020** | 0.36 | 0.90 | 0.92 | 0.91 | 0.92 | 1 |  |  |  |
| **Feb 2020** | 0.35 | 0.83 | 0.87 | 0.88 | 0.91 | 0.91 | 1 |  |  |
| **Mar 2020** | 0.44 | 0.88 | 0.91 | 0.90 | 0.92 | 0.92 | 0.90 | 1 |  |
| **Apr 2020** | 0.45 | 0.84 | 0.89 | 0.90 | 0.90 | 0.91 | 0.89 | 0.95 | 1 |

